# Bridging Theory and Behavior for Healthcare Accessibility Modeling: A Mobility-Driven Revision of the E2SFCA

**DOI:** 10.1101/2025.09.10.25334268

**Authors:** Ali Khosravi Kazazi, Zhenlong Li

## Abstract

Proximity to a healthcare supplier does not necessarily equate to meaningful access to care. Traditional healthcare accessibility models, particularly the Enhanced Two-Step Floating Catchment Area (E2SFCA) method, rely heavily on assumptions of proximity-driven behavior, fixed catchment sizes, and uniform distance decay. These simplifications often overlook the complexities of real-world healthcare-seeking behavior. This study examines the assumptions of the E2SFCA framework by integrating large-scale human mobility data, which captures anonymized, real-world visitation patterns between neighborhoods and hospitals across Pennsylvania. We revise the E2SFCA model through two key innovations: 1) replacing static catchment thresholds with dynamic, visit-weighted boundaries derived from observed travel behavior, and 2) estimating hospital-specific distance decay functions that better reflect heterogeneous patterns of attraction. These refinements result in accessibility metrics that align more closely with empirical realities. Compared to the traditional model, the revised E2SFCA demonstrates a more meaningful relationship with real-world outcomes, showing stronger correlation with household income and a more plausible negative association with poor or fair health status. Additionally, the revised model reveals significantly greater inequality in access, with the Gini coefficient nearly doubling (0.63 vs. 0.31), and shows that 19% of the population accounts for 63% of accessibility, exposing disparities masked by proximity-based approaches. By integrating human mobility data in spatial accessibility modeling, this study offers a more realistic, equitable, and policy-relevant framework for evaluating healthcare access.

## 1. Introduction

Accessibility, defined as the ease of reaching services from a location, is critical for achieving equitable public health outcomes (Wang, 2012). The World Health Organization (WHO) highlights healthcare accessibility as a key factor in reducing disparities(World Health Organization, n.d.), requiring accurate measurement to identify underserved populations. Researchers have developed diverse methods to evaluate access, ranging from simple metrics like provider-to-patient ratios to advanced models simulating individual rational choices (Saxon & Snow, 2020). These efforts also distinguish between different types of accessibility, such as objective measures and subjective, perception-based experiences, while acknowledging barriers including socioeconomic disadvantage, transportation gaps, and systemic inequities. (Negm et al., 2025; Tzenios, 2019).

A widely used approach for measuring healthcare accessibility is the Floating Catchment Area (FCA) method, which aims to balance conceptual simplicity and practical implementation. The Enhanced Two-Step Floating Catchment Area (E2SFCA) method (Luo & Qi, 2009a) builds on earlier models by incorporating supply, demand, and distance decay, the principle that the likelihood of visiting a provider decreases as travel distance increases. While E2SFCA has proven effective in identifying underserved areas, it also has notable limitations. First, it relies on fixed distance thresholds to define catchment areas, leading to abrupt discontinuities in accessibility scores. For instance, a resident living just beyond a mile threshold may be excluded from accessing a nearby supplier, despite its practical proximity. Second, the method is susceptible to edge effects, potentially misclassifying individuals near geographic boundaries if nearby healthcare facilities fall outside the defined catchment. Third, it assumes uniform travel behavior across populations, overlooking important variations such as rural residents who typically travel longer distances, or individuals with disabilities who may face greater mobility constraints (Drake et al., 2021). The model also assumes people prioritize proximity, which may oversimplify real-world healthcare decisions. Patients often bypass closer facilities due to insurance networks, referrals, or preferences for specialized care (Victoor et al., 2012). For instance, a cancer patient might travel long distances to a specialty hospital, defying proximity-based assumptions.

To address these gaps, scholars such as Wang (2012) advocate for grounding accessibility models in empirical data (Wang, 2012). Emerging mobility datasets such as anonymized SafeGraph mobile location data (now Advan patterns) provide valuable insights into actual travel behavior. SafeGraph aggregates visits to facilities from millions of mobile devices, capturing place visitation patterns such as cross-regional healthcare utilization (Pan et al., 2018). While challenges remain such as sampling biases that may underrepresent low-income or older populations (Li et al., 2024), such data help bridge the gap between theoretical models and real-world dynamics

This study aims to address the limitations of conventional FCA methods by integrating population mobility data into the E2SFCA framework. We revise catchment areas based on observed visitation patterns, replacing fixed distance thresholds with dynamic boundaries that reflect actual patient travel behaviors. Additionally, we refine the distance decay component by deriving hospital-specific decay functions from empirical data, recognizing, for example, that a regional trauma center may draw patients from greater distances, while a neighborhood hospital serves a more localized population. These enhancements are synthesized into a revised E2SFCA model that evaluates healthcare accessibility through a dual lens: one rooted in spatial theory, the other grounded in real-world mobility patterns.

## 2. Related Works

Access to healthcare is broadly defined as the “fit” between patient needs and the availability of services (Gulliford et al., 2002). Seminal frameworks describe multiple dimensions of access -availability, accessibility (geographic), affordability, accommodation, and acceptability-that together determine how well services match patient needs (Israel, 2016). Levesque et al. (2013) similarly conceptualized access through five abilities of populations to seek care: approachability (awareness of services), acceptability (cultural/social fit), availability and accommodation (physical reach and organization), affordability (economic cost), and appropriateness (fit of services to needs) (Levesque et al., 2013). These models highlight that spatial barriers (distance, travel time), financial barriers (cost, insurance), and perceived barriers (trust, cultural congruence, knowledge) all influence access. In sum, healthcare accessibility is multidimensional, reflecting service supply relative to demand as well as personal, social, and economic factors.

Early measures of geographic access were relatively simple, often relying on provider-to-population ratios-such as the number of providers per capita in an area, were widely used (e.g., U.S. HPSA scores) (Donohoe et al., 2016). Distance or travel-time measures (e.g., to nearest provider) also became common. However, these one-dimensional metrics ignore spatial distribution of supply and demand. To capture both, gravity-based models were introduced. Joseph & Bantock (1984) proposed an accessibility model that integrates provider capacity and population demand with a distance-decay function (Joseph & Bantock, 1982). Building on this, Luo and Wang (2003) developed the Two-Step Floating Catchment Area (2SFCA) method: first computing a provider-to-population ratio within a travel-time catchment around each provider, then summing these ratios for each population location’s catchment (Luo & Wang, 2003). The 2SFCA thus yields a local supply-demand ratio that accounts for overlapping service areas. Numerous enhancements followed. Luo & Qi (2009) introduced the Enhanced 2SFCA (E2SFCA) by applying distance-decay weights within catchments, giving nearer populations more weight (Luo & Qi, 2009b). Other studies have implemented continuous distance-decay kernels, variable catchment sizes, multi-modal travel, and other refinements(Guagliardo et al., 2004; Langford et al., 2016; Ni et al., 2015). These evolving methods, from simple ratios and gravity models to floating catchment approaches (e.g., 2SFCA, E2SFCA), aim to better represent the geographic reality of healthcare access.

Despite its advances, the E2SFCA (and related FCA) framework has known limitations. (1) Fixed catchment sizes: Standard 2SFCA/E2SFCA typically use one travel-time radius (e.g., 30–60 minutes) for all locations. This ignores that urban populations may accept short travel while rural populations tolerate longer trips. (2) Edge effects: Restricting analysis to administrative boundaries can bias results because people near borders may use providers across the border, but these are excluded, leading to artificial “access deserts” (Donohoe et al., 2016). (3) Uniform travel assumptions: These methods assume homogeneous travel behavior (same speed, mode) for all individuals. In reality, mobility varies by age, car ownership, disability, etc., which the models do not capture. (4) Dichotomous accessibility: Traditional 2SFCA (and even E2SFCA with stepwise weights) treats providers within the threshold as fully accessible and those outside as inaccessible(Luo & Qi, 2009a). This sharp cut-off is unrealistic because in reality access declines gradually with distance. (5) Proximity prioritization: Implicitly, these models assume individuals use the nearest available providers first, filling up capacity in distance order, which may not reflect personal preferences or provider choice behavior. (6) Overcounting demand: By assigning full population demand to every facility within range, 2SFCA can overestimate demand if individuals are counted at multiple providers simultaneously. In short, E2SFCA simplifies human behavior and spatial context (fixed zones, equal weights), leading to edge effects and untested assumptions about travel and choice(Donohoe et al., 2016).

Recent years have seen growing use of real-world mobility data to study healthcare and service access. Data sources include anonymized mobile-phone location datasets (SafeGraph, Cuebiq, Google, etc.) and call detail records (CDRs) from telecom providers. These capture where people actually travel and which facilities they visit. For example, Wang et al. (2021) used SafeGraph data to analyze patterns of healthcare visits during the COVID-19 pandemic(Wang et al., 2021). They clustered census block groups by temporal trends in visits and linked declines to demographic factors, illustrating how mobility data reveal access disparities in practice. More broadly, Swanson & Guikema (2024) developed methods using cellphone “location-based services” data to assess community-level loss of access to essential services (including healthcare) after disasters (Swanson & Guikema, 2024). These studies demonstrate strengths of mobility data: large-scale, high-frequency observations of actual travel behavior and utilization.

A few emerging studies have begun to integrate mobility into spatial accessibility models. Chen et al. (2024) propose a Generalized Flow-based 2SFCA (GF2SFCA) that explicitly uses observed patient flows (from mobility data) to inform the model (Chen et al., 2024). In GF2SFCA, hospital catchment sizes, distance-decay rates, and attractiveness weights are data-driven: for example, hospitals’ “global popularity” and “local preference” indices are computed from actual visitation counts and travel distances. In a Wuhan case study, this approach produced more realistic accessibility maps than conventional 2SFCA and showed robustness to data uncertainties(Chen et al., 2024). Though focused on hospital access, the principle applies more broadly. Other work (e.g., in urban parks and food access) similarly uses travel-behavior data to adjust catchment radius and decay functions. Overall, these hybrid models improve realism by calibrating FCA parameters with real mobility, moving beyond arbitrary assumptions.

The literature reveals at least two notable gaps in current healthcare accessibility research. First, the integration of actual mobility data into FCA models remains limited. While recent methodological advances like the GF2SFCA (Chen et al., 2024) have been proposed, hybrid approaches that combine empirical travel data with FCA methods are still rare. Second, there is a lack of systematic validation of the core assumptions underlying the E2SFCA model, such as fixed travel thresholds, uniform distance decay, and proximity-based behavior, against observed patient travel patterns. Researchers have pointed out that the scarcity of patient travel data constrains this validation (Donohoe et al., 2016). Consequently, the behavioral assumptions foundational to E2SFCA remain largely untested and unverified in empirical research.

The literature also highlights the importance of grounding healthcare accessibility models in actual mobility patterns. In the context of Pennsylvania, prior research illustrates the stakes: Drake et al. (2021) found that basic measures such as straight-line distance or provider-to-population ratios failed to identify 30–52% of census tracts that the E2SFCA model classified as underserved (Drake et al., 2021), emphasizing the need for more sophisticated spatial approaches. Building on these insights, our study employs SafeGraph mobility data to empirically revise key E2SFCA parameters. Specifically, we analyze observed visit flows to Pennsylvania hospitals to define catchment boundaries and assign distance decay weights that reflect real-world travel behavior. This integration directly addresses known limitations of the E2SFCA model, for example, replacing uniform catchment thresholds with dynamic radius that reflect actual willingness to travel, and tailoring decay functions to match empirically observed declines in visitation with distance. The resulting model, which integrates the theoretical strengths of FCA accessibility with empirical mobility data, represents a novel contribution.

## 3. Methodology

This study systematically evaluates the core assumptions of the E2SFCA method by comparing them against empirical human mobility patterns to assess their validity in real-world context. The E2SFCA method is built on four main assumptions: 1) both suppliers and demanders operate within bounded geographic catchment areas (Luo & Qi, 2009a); 2) these catchments are fixed and of equal size across all locations (Stacherl & Sauzet, 2023); 3) within these catchments, individuals’ willingness to access services decreases with increasing distance (distance decay) (Delamater, 2013); and 4) this distance decay is identical for all suppliers and demanders, regardless of local variation (McGrail, 2012). For each assumption, we follow a two-step process. First, we evaluate its alignment with empirical data by comparing the theoretical premise to observed mobility patterns, including visitation flows and distance decay trends. Second, based on this assessment, we either refine or revise the assumption as needed. If an assumption aligns with empirical evidence, we enhance its precision (e.g., replacing fixed catchment sizes with thresholds derived from observed mobility data). If it diverges from observed behavior, we discard or reformulate it (e.g., substituting uniform distance decay with variable, context-specific functions). By anchoring each assumption in real-world data, this study bridges the gap between spatial accessibility theory and observed healthcare-seeking behavior.

### 3.1 Data and study setting

This study focuses on Pennsylvania, U.S. The state’s geographic diversity introduces substantial variation in healthcare infrastructure and physical accessibility, shaped by natural barriers, population density gradients, and uneven hospital distribution. These spatial disparities intersect with demographic and structural inequities: rural communities face aging populations and mobility constraints, while wealth disparities and environmental health risks disproportionately burden underserved urban and peri-urban neighborhoods. Together, these dynamics make Pennsylvania a compelling case study for analyzing spatial accessibility in a way that reflects broader national trends.

The analysis integrates multiple data sources across three spatial scales: block groups (n = 9,740), counties (n = 67), and hospitals (n = 191). Demographic and socioeconomic data, including population and mean household income, were obtained from the U.S. Census Bureau’s American Survey (ACS) 5-Year Estimates for 2018-2022. These data provide the demand-side context for block groups and support county-level aggregation for validation against public health outcomes (U.S. Census Bureau, 2023). Hospital data were acquired from the Commonwealth of Pennsylvania Department of Health, including facility names, addresses, and the number of licensed beds as a proxy for service capacity (Commonwealth of Pennsylvania Department of Health, 2024). The dataset encompasses a full spectrum of hospital types, including general acute care, specialty, and federal hospitals. Health outcome data, specifically the percentage of the population reporting poor or fair health, were sourced from the County Health Rankings and Roadmaps (University of Wisconsin Population Health Institute, 2025), allowing for validation of accessibility scores at the county level.

To capture empirical travel behavior, the study utilizes SafeGraph’s (now Advan patterns) anonymized human mobility dataset, which includes over 3.5 million hospital visits across Pennsylvania during calendar year 2023. Visits were aggregated at the census block group level and linked to hospitals using address matching. To accurately link mobility visits to hospitals, we implemented a structured address-matching process. The hospital dataset provided facility names and county information, which we used to query the Google Maps API and obtain standardized addresses. We then used the Placekey platform (Placekey, 2024) to find the unique Placekey for each hospital, which separates spatial location ("where") from descriptive attributes ("what"). To ensure consistent spatial referencing, we extracted the “where” component of each Placekey and compared it to those in the mobility dataset. For each match, we applied fuzzy string matching (Wu, 2016) to hospital names to identify the most likely corresponding record. This approach enabled reliable linkage between hospital locations and mobility data, even when address formats or facility names varied slightly. In total, 191 hospitals were successfully matched and included in the final analysis. Geodesic distances, measured in miles between centroids of origins and destinations, were used to estimate travel distances in all modeling steps.

### 3.2. Evaluate the bounded catchment areas assumption

A central methodological debate in spatial accessibility modeling concerns whether to constrain patient–provider interactions within predefined catchment areas or to adopt an open, boundary-free approach. Catchment-based models, such as the E2SFCA, impose geographic limits (typically based on travel time or distance), around healthcare providers and their surrounding populations. While this structure simplifies computation and aligns with administrative planning, it risks oversimplifying real-world mobility, as patients may bypass nearby facilities in favor of more specialized or preferred providers farther away. In contrast, open models remove arbitrary thresholds and can capture long-distance travel patterns, but they risk overestimating accessibility by assuming that all populations, including those in remote areas, have equal access to distant hospitals, potentially introducing noise from unrealistic interactions.

This assumption test empirically evaluates whether catchment areas reflect actual human mobility behaviors and seeks to clarify whether spatial accessibility frameworks should impose bounded catchment areas or adopt a more flexible, boundary-free approach grounded in empirical mobility data.

From the supplier (hospital) perspective, we compute:

1. Hospital coverage ratio:

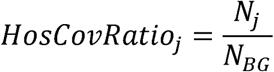

where *N_j_* is the number of unique block groups visiting hospital *J* and *N_BG_* is the total number of block groups.

2. Relative visit-weighted distance for hospitals (HosRWD)

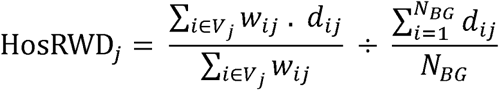

where *w_ij_* is the number of visits from block group *i* to hospital *j*, *d_ij_* is the distance between them and *V_j_* is set of block groups with non-zero visits to hospital *j*.

From the demand (population at block group level) perspective, we compute:

1. Hospital Choice Ratio (PopChoiceRatio)

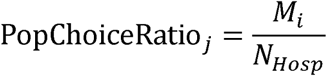

where *M*i is the number of unique hospitals visited by block group *i* and *N_Hosp_* is the total number of hospitals

2. Relative visit-weighted distance for block groups (PopRWD)

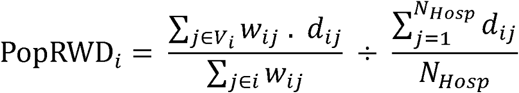

where *V_i_* is set of hospitals visited by block group *i*.

Lower values in these metrics (especially the relative distance ratios) indicate more localized, catchment-like behavior, supporting the use of geographic boundaries in accessibility models. Higher values suggest broader or more dispersed travel behavior, consistent with an open, boundary-free model.

### 3.3. Evaluate the fixed catchment sizes assumption

A key critique of fixed catchment sizes is their failure to account for variations in travel behavior, particularly the tendency of rural populations to travel longer distances for care compared to urban residents. Numerous studies argue that catchment boundaries should reflect observed mobility patterns rather than rely on arbitrary thresholds. To address this limitation, our study introduces a visit-weighted average distance metric for both population origins and hospital destinations.

For each block group *i*, the visit-weighted average distance to utilized hospitals is calculated as:

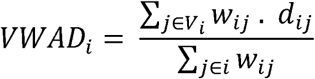

where *w_ij_* is the number of visits from block group *i* to hospital *j*, and *d_ij_* is the distance between them.

Similarly for each hospital *j* the average distance from its visitors is:

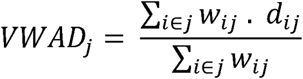

This metric captures how far individuals actually travel by weighting each distance according to the number of visits to a given hospital.

This approach offers three distinct advantages. First, it enhances behavioral realism by prioritizing observed travel behavior over theoretical assumptions. Second, it incorporates visit intensity, ensuring that hospitals frequently visited by patients exert a proportionally greater influence on the calculated average, thus capturing disparities in healthcare utilization. Third, it accounts for heterogeneity in both provider and population behavior, recognizing that certain hospitals (e.g., specialized or regional centers) may attract patients from greater distances, while some communities routinely travel farther for care.

### 3.4. Distance decay consideration

The E2SFCA method incorporates distance decay in two sequential steps: 1) hospitals attract fewer visitors as the distance to surrounding neighborhoods increases, and 2) residents reduce their utilization of hospitals as travel distance grows. This dual-decay framework assumes a symmetrical decline in interaction likelihood between supply and demand as spatial separation increases. To evaluate this assumption, we fit both power-law and linear (negative exponential in log-linear form) distance decay models to empirical hospital–neighborhood visit data. The power-law model takes the form 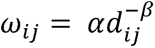 while the exponential model is expressed as 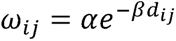. For each hospital, we estimated model parameters and assessed fit quality using two criteria: coefficient of determination (R²) and Akaike Information Criterion (AIC). We then classified the best-fitting model using a systematic rule-based approach.

A hospital was classified as following a power-law decay if the power-law model outperformed the linear model on both R² and AIC metrics, and its distance coefficient was significantly negative (*β*,< 0, *p* < 0.05). Conversely, it was classified as following a linear decay if the linear model showed superior fit by the same criteria and also had a significantly negative coefficient. If neither model met these conditions, i.e., if the better-fitting model lacked a statistically significant negative coefficient, the hospital was categorized as exhibiting no clear decay.

### 3.5. Evaluate the uniform distance decay assumption

The E2SFCA method assumes a uniform distance decay function applies identically to all suppliers and neighbors, disregarding potential variability in spatial interaction behavior. To test this assumption, we estimate hospital-specific decay parameters using a power-law model (*W*(*d*) - *d*^-*β*^). For each hospital, we fit the model to its visitation data to derive a unique decay parameter (*β*), reflecting how rapidly visitation frequency declines with distance. This approach allows us to quantify heterogeneity through comparing decay rates across suppliers and evaluate uniformity to assess whether a single decay parameter is sufficient. This test is expected to provide new insights on whether the uniform decay assumption aligns with empirical patterns or obscures critical variations in healthcare-seeking behavior, advocating for context-sensitive decay functions in spatial accessibility modeling.

### 3.6. Development of revised method

Following the independent evaluation of the four core assumptions underlying the E2SFCA method, we develop a revised spatial accessibility framework that integrates the methodological insights derived from the preceding analyses. The proposed framework incorporates key modifications to the original E2SFCA structure. These adjustments are designed to enhance the model’s capacity to represent observed patterns of healthcare-seeking behavior and spatial interaction.

## 4. Results and Discussion

The results highlight how traditional spatial accessibility metrics that are rooted in assumptions of proximity-driven behavior and uniform distance decay can misrepresent actual patterns of healthcare utilization. By recalibrating the E2SFCA model using empirically derived dynamic catchment areas and variable distance decay parameters, we reveal more context-sensitive disparities in spatial access. These findings demonstrate that mobility-informed refinements yield a more realistic and equitable representation of healthcare accessibility.

### 4.1. Bounded catchment areas

This section empirically evaluates a core assumption underpinning spatial accessibility models like the E2SFCA method: that healthcare interactions are geographically constrained and typically occur within bounded catchment areas. While this assumption is often taken for granted in the design of such models, few studies have systematically quantified the extent to which real-world healthcare interactions align with this theoretical framework.

As shown in Table 1, our analysis reveals a pronounced localization in hospital–patient interactions. From the hospital perspective, the average visit-weighted distance to patients (HosRWD) is only 33.53 miles, in contrast to the 130.9-mile average distance across all block groups. On average, each hospital serves only 340 out of 9,740 block groups, or about 3.5% of the possible population base. This indicates a sharply bounded spatial reach for most hospitals. From the block group perspective, the average visit-weighted distance to utilized hospitals (PopRWD) is just 14.7 miles, compared to 130.9 miles if all hospitals were considered equally. Furthermore, each block group engages with only 5 out of 191 hospitals on average (2.6%), reinforcing the conclusion that residents overwhelmingly seek care from a small, nearby subset of providers.

**Table 1.**
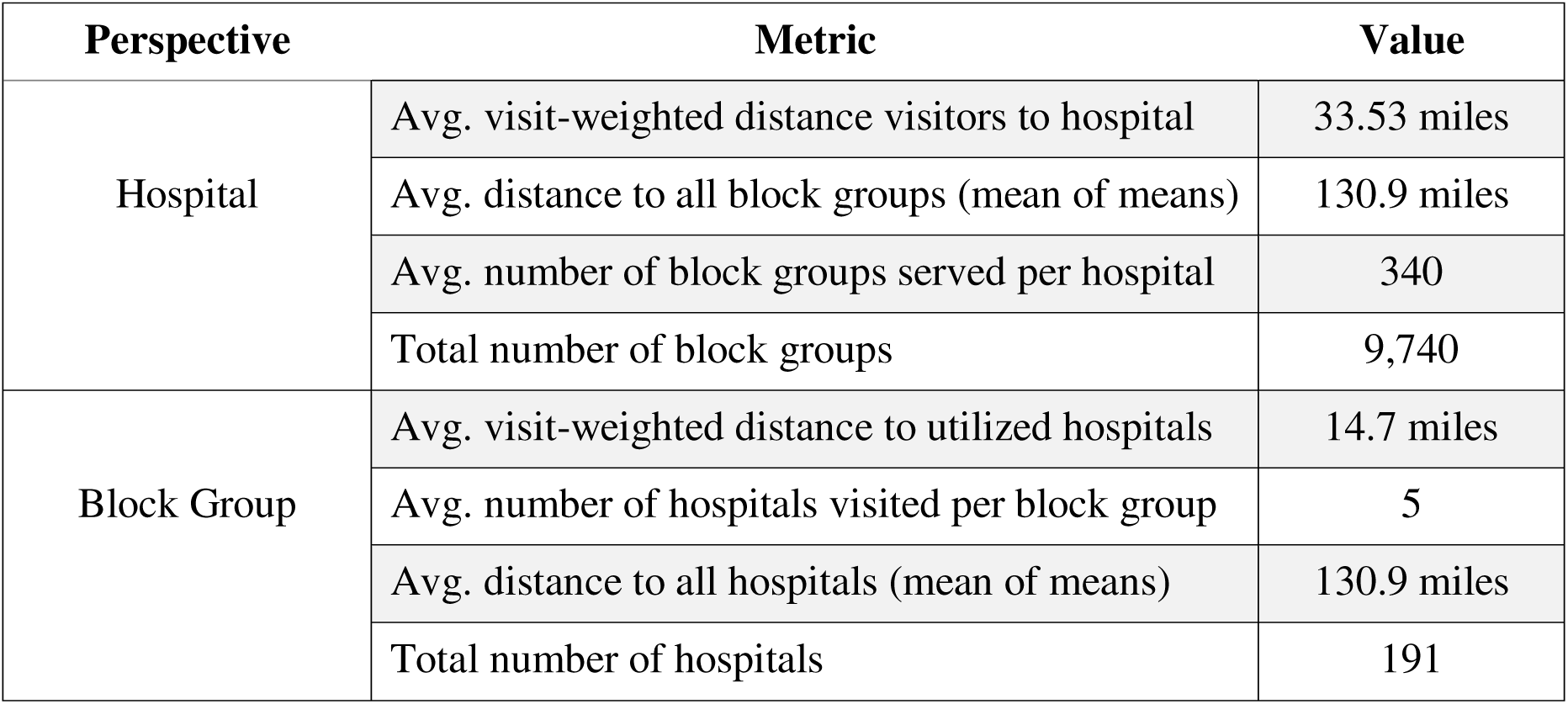
Summary of interaction patterns between hospitals and block groups.

Although the original E2SFCA method implicitly assumes localized interactions via predefined catchments and distance decay functions, it does not specify whether these assumptions match empirical behavior. The significant differences between actual travel distances and theoretical full-network distances validate that spatial interaction is indeed bounded in practice. This spatial concentration of interactions justifies the continued use of catchment areas in spatial accessibility models.

### 4.2. Fixed catchment sizes

While the previous section demonstrated that patient-provider interactions are geographically bounded, this section focuses on how those bounds are represented in accessibility models. Specifically, it critiques the common use of fixed-distance catchment sizes, showing that actual travel behavior varies significantly across contexts and cannot be captured by a one-size-fits-all threshold.

To more accurately reflect real-world travel behavior, we employed a visit-weighted average distance metric, which adjusts travel distances based on the frequency of visits to each hospital. Unlike arbitrary static thresholds (e.g., a fixed 30-mile catchment), this metric captures the actual spatial extent of interactions, offering a more behaviorally grounded approach to defining access. The average VWAD for all hospitals was 33.5 miles (median = 28.9 miles), while residents traveled an average of 14.7 miles (median = 11.14 miles) to access care. These findings suggest that a one-size-fits-all catchment may either overestimate or underestimate access depending on local conditions, provider type, and population characteristics.

Figure 1 illustrates the variability of catchment sizes by mapping dynamic catchment areas for hospitals, where the spatial reach expands or contracts based on actual visitation patterns. Figure 2 shows catchment sizes for block groups. The underlying data reveal similarly uneven patterns of hospital utilization. These findings suggest that visit-weighted distance metrics offer a more flexible and empirically grounded alternative to fixed catchment sizes, allowing spatial accessibility models to better reflect how far people actually travel for care.

**Figure 1.**
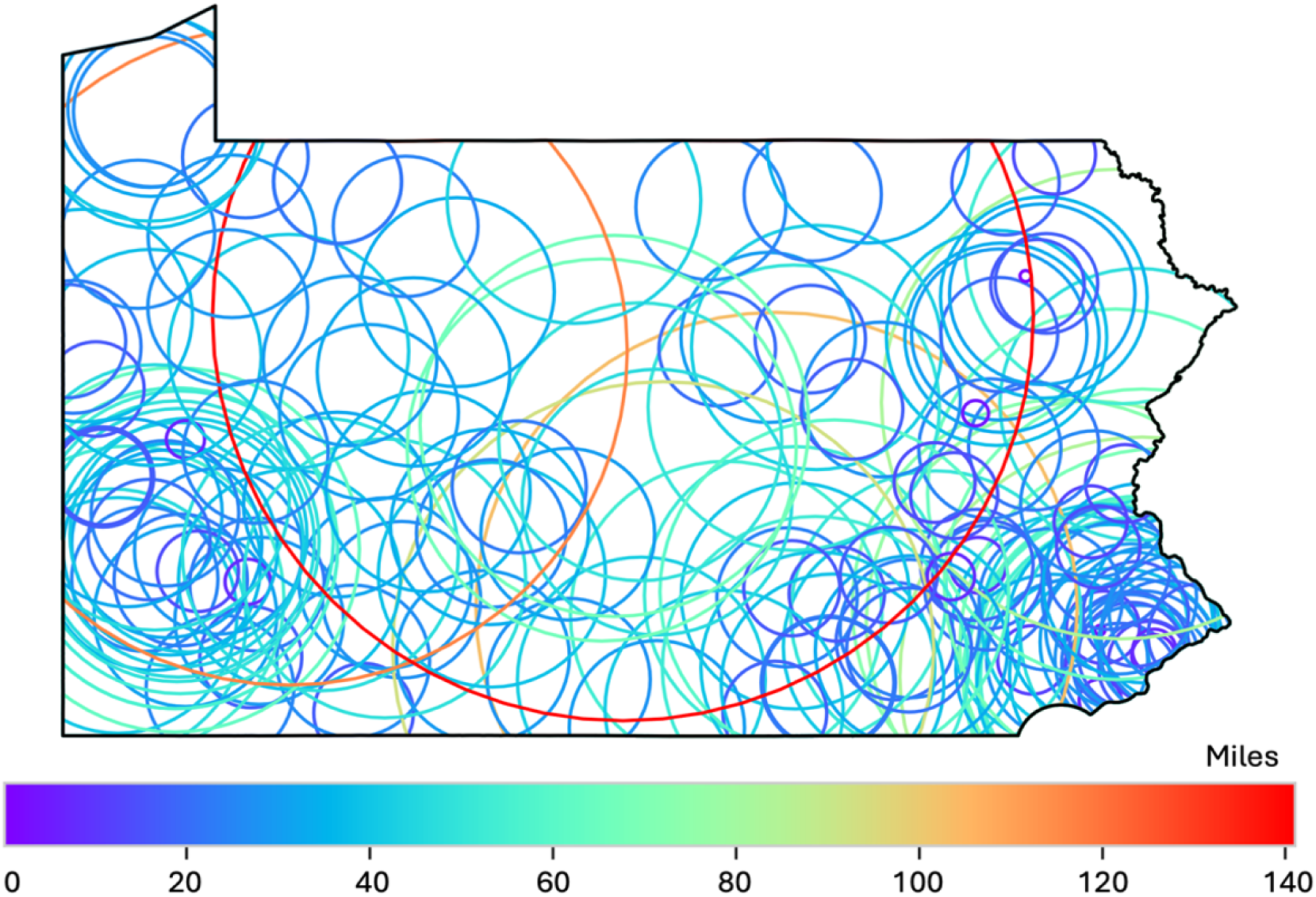
Hospitals’ dynamic catchment area

**Figure 2.**
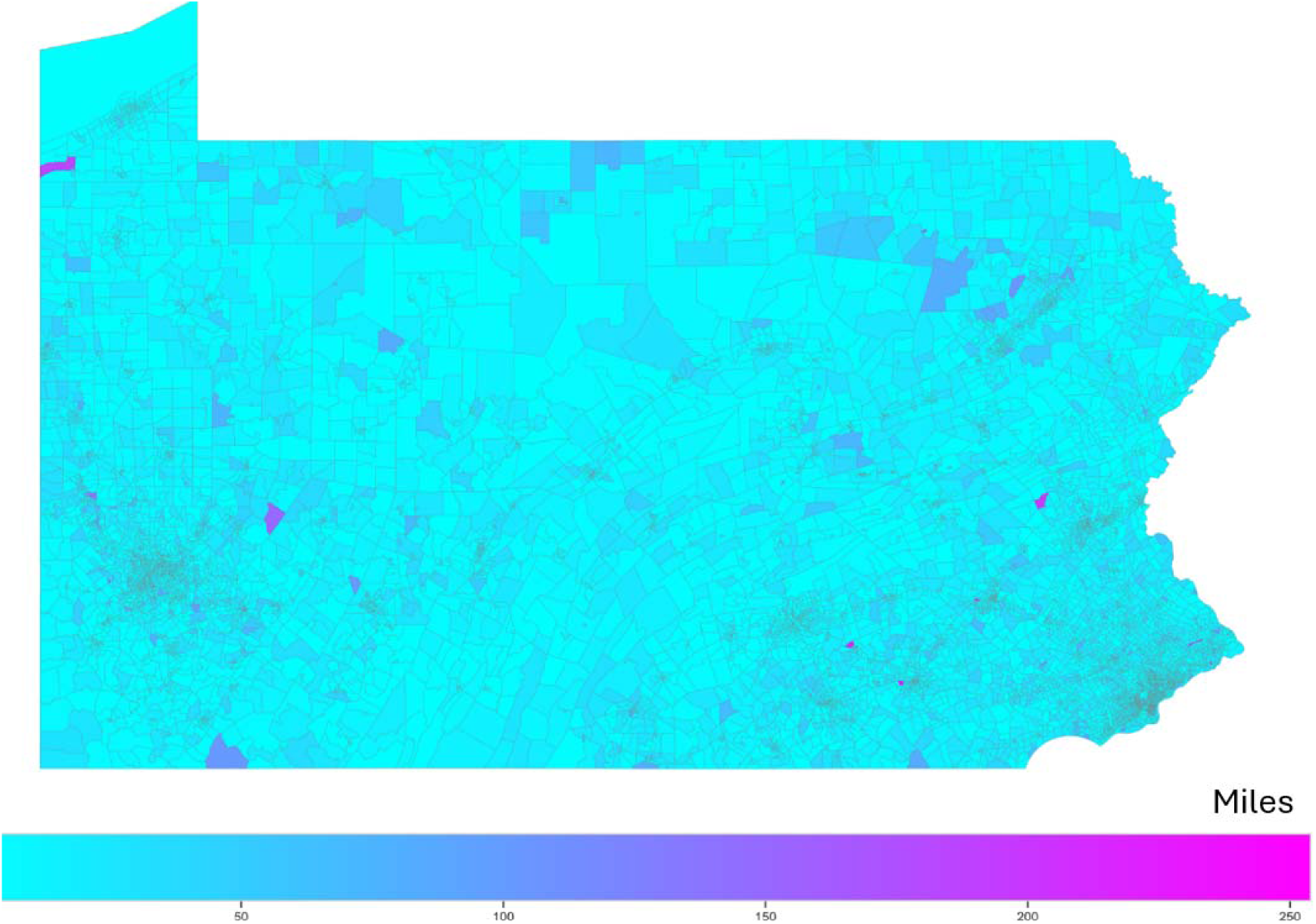
Block groups’ dynamic catchment sizes

### 4.3. Distance decay consideration

The analysis of distance decay reveals a critical asymmetry in how spatial proximity influences healthcare interactions from the perspectives of hospitals and block groups. Using model classification based on both statistical fit (R², AIC) and coefficient significance, we assessed whether hospital–neighborhood visitation patterns conformed to power-law, linear (log-linear), or no discernible distance decay behavior. Table 2 summarizes the distribution of decay classifications.

**Table 2.** Model fit comparison for distance decay patterns.

| Model | Hospitals (%) | Block Groups (%) |
| --- | --- | --- |
| Power-Law | 80.1 | 15 |
| Linear | 0 | 1 |
| No Decay | 19.9 | 65 |
| No Data | 0 | 19 |

From the hospital perspective, results support the expected pattern of distance-based interaction: for 80.1% of hospitals, the power-law model provided the best fit, indicating a gradual, nonlinear decline in patient volume with increasing distance. An additional 19.9% of hospitals exhibited no statistically significant decay, while none were best described by a linear model. This suggests that, in general, hospitals attract fewer visitors as distance increases, consistent with the spatial assumptions embedded in the E2SFCA framework.

In contrast, the block group perspective reveals a different pattern. For 65% of block groups, visitation behavior showed no significant relationship with distance i.e., proximity to hospitals did not predict utilization. Only 15% were best fit by a power-law decay model, less than 1% by a linear model, and approximately 19% lacked sufficient data for analysis. These findings indicate that, for most neighborhoods, healthcare-seeking behavior is not driven by geographic accessibility alone. Non-spatial factors such as insurance coverage, provider reputation, referral networks, and perceived quality likely play a dominant role in shaping hospital choice.

Figure 3a illustrates a hospital with a clear power-law decay pattern, where visitation drops sharply beyond half a mile. In contrast, Figure 3b depicts a hospital with no evident decay, where visit frequency remains constant across distance.

**Figure 3.**
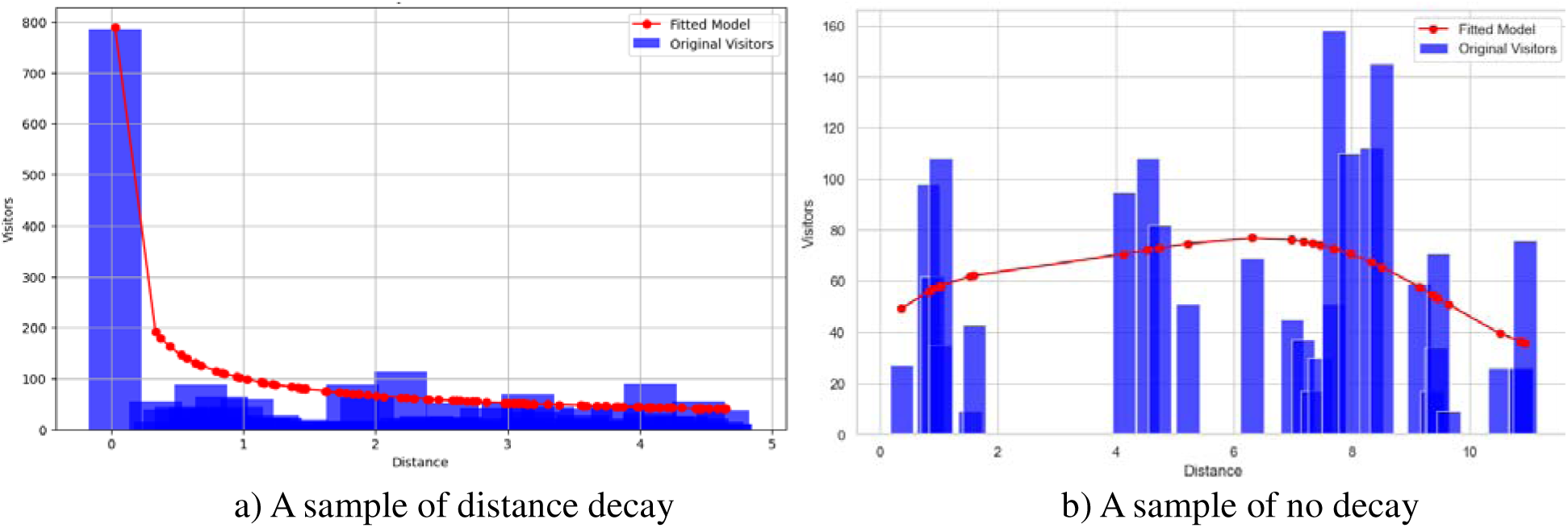
Two samples of the distribution of visits over distance with fitted power-law model

Together, these results indicate the limitation of the E2SFCA’s assumption of symmetrical distance decay, which posits that both suppliers and demanders reduce interaction likelihood as distance increases. While hospitals largely conform to this assumption, patient behavior does not. This asymmetry suggests that distance decay should be applied selectively, primarily to supplier catchments, and highlights the limitations of purely spatial models in capturing the full complexity of healthcare access. To consider this realism, future accessibility frameworks should account for non-spatial drivers of patient choice and reconsider the application of decay functions across both supply and demand dimensions.

### 4.4. Uniform distance decay

To evaluate the assumption of uniform distance decay across hospitals, we modeled visitation patterns using a power-law function: visitors =*α.*d^-^*^β^* where *α* represents baseline visitation (i.e., interaction at short distances), and β denotes the decay rate. Each hospital was fitted with its own set of parameters, allowing us to capture provider-specific distance decay behavior.

The median decay rate β across hospitals was –0.26, indicating a relatively gradual decline in visitation with increasing distance for most providers. The mean β at –0.24, closely aligned with the median, suggesting a relatively symmetric distribution with limited influence from outliers. This pattern points to greater consistency in distance decay behavior across providers than previously observed. In contrast, the baseline visitation parameter (α) exhibited more variation. The median α was 38.8, reflecting strong local interaction for a typical hospital, while the mean α was higher at 56.1, indicating a moderate right-skew driven by hospitals experiencing unusually high visitation from nearby populations. This wide variability in both decay and baseline interaction reveals the limitations of using a uniform distance decay function across all providers. These findings support the need for hospital-specific decay parameters to more accurately reflect the diverse spatial behaviors in healthcare utilization.

### 4.5. Revised E2SFCA method

Building on the empirical evaluation of the four foundational assumptions of the E2SFCA method, we developed and implemented a revised spatial accessibility model that reflects observed patterns in healthcare-seeking behavior. The two steps of the revised method are:

***Step 1:*** Estimate the distance-decay-adjusted supply-to-demand ratio for provider *j* using an empirically fitted power-law decay function.

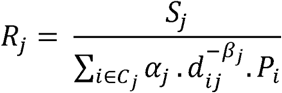

where *s_j_* is supply at provider *j*, *P_i_* is population at location *i*, *d_ij_* is the distance between demander *i* and supplier *j*, *α_j_* and *β_j_* are provider-specific decay parameters and *C_j_* is dynamic catchment for provider *j* with a radius equal to *VWAD_j_*.

***Step 2:*** For each population location *i*, define its dynamic catchment *C_j_*, based on its

*VWAD_i_*. Then, aggregate all accessible providers within this radius to compute the accessibility score.

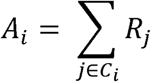

### 4.6. Accessibility maps

The study concludes with a comparative analysis of healthcare accessibility in Pennsylvania, contrasting the original E2SFCA method with default parameters from Darke et al. (2021) with the revised E2SFCA framework empirically calibrated using human mobility data. (Drake et al., 2021). To ensure comparability, the accessibility scores from both approaches were normalized using a min–max scaler, rescaling values between 0 and 1. Figure 4 presents accessibility scores derived from the original E2SFCA method. As expected, these scores exhibit a proximity-driven pattern: neighborhoods located near hospitals receive uniformly high accessibility ratings, while access steadily declines with increasing distance.

**Fig 4.**
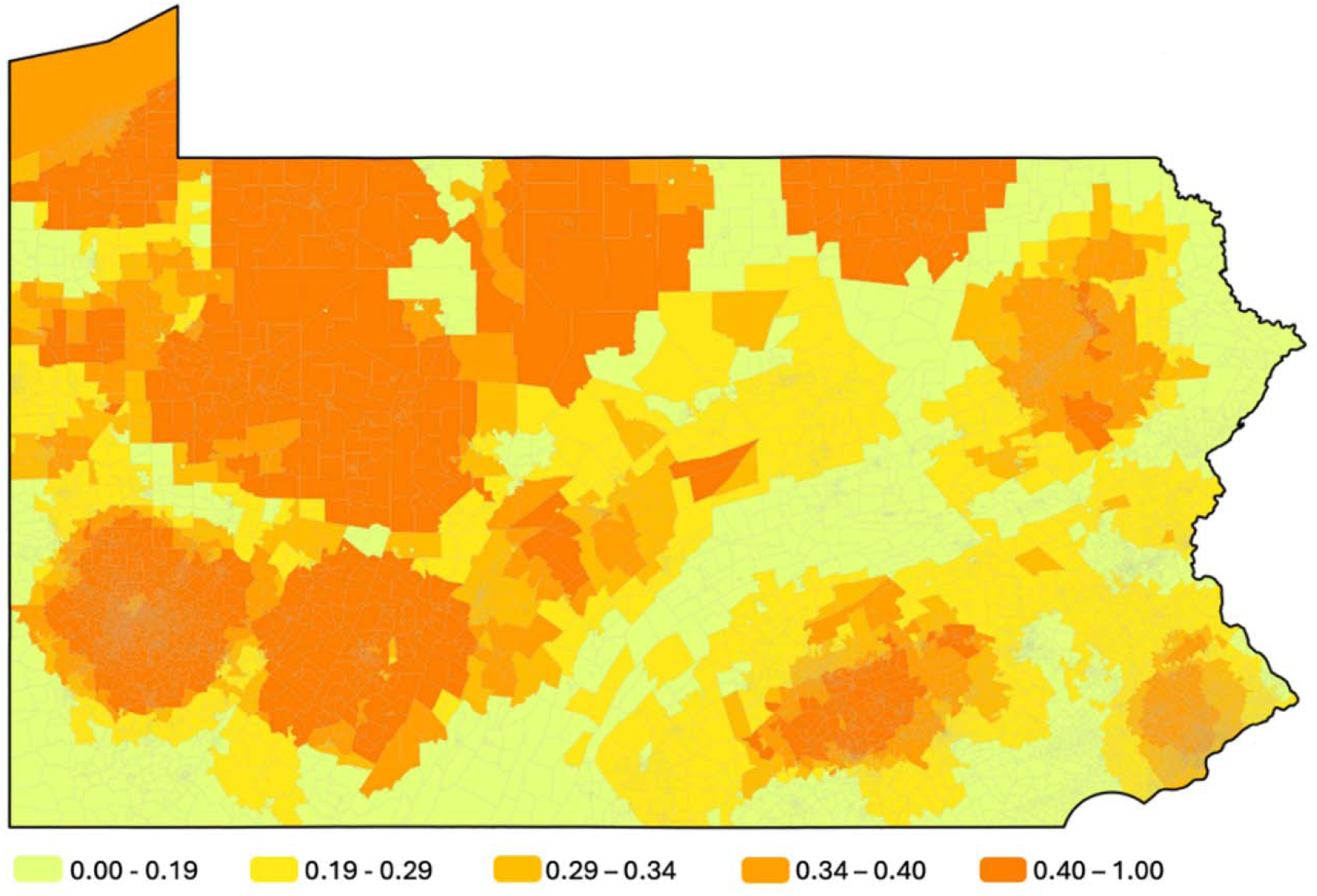
Accessibility map based on E2SFCA and default parameters

This pattern reflects the underlying assumptions of fixed catchment sizes and uniform distance decay. In contrast, Figure 5 maps accessibility using the revised E2SFCA model, which incorporates dynamic catchment areas, hospital-specific distance decay rates, and empirically derived attractiveness indices. This approach produces a more nuanced accessibility landscape, revealing disparities that are obscured by the traditional model, such as hospitals with high regional pull or underserved areas near densely located but overburdened providers. By visualizing and comparing both approaches, this analysis demonstrates how empirically grounded refinements can improve the realism and equity of spatial accessibility assessments.

**Fig 5.**
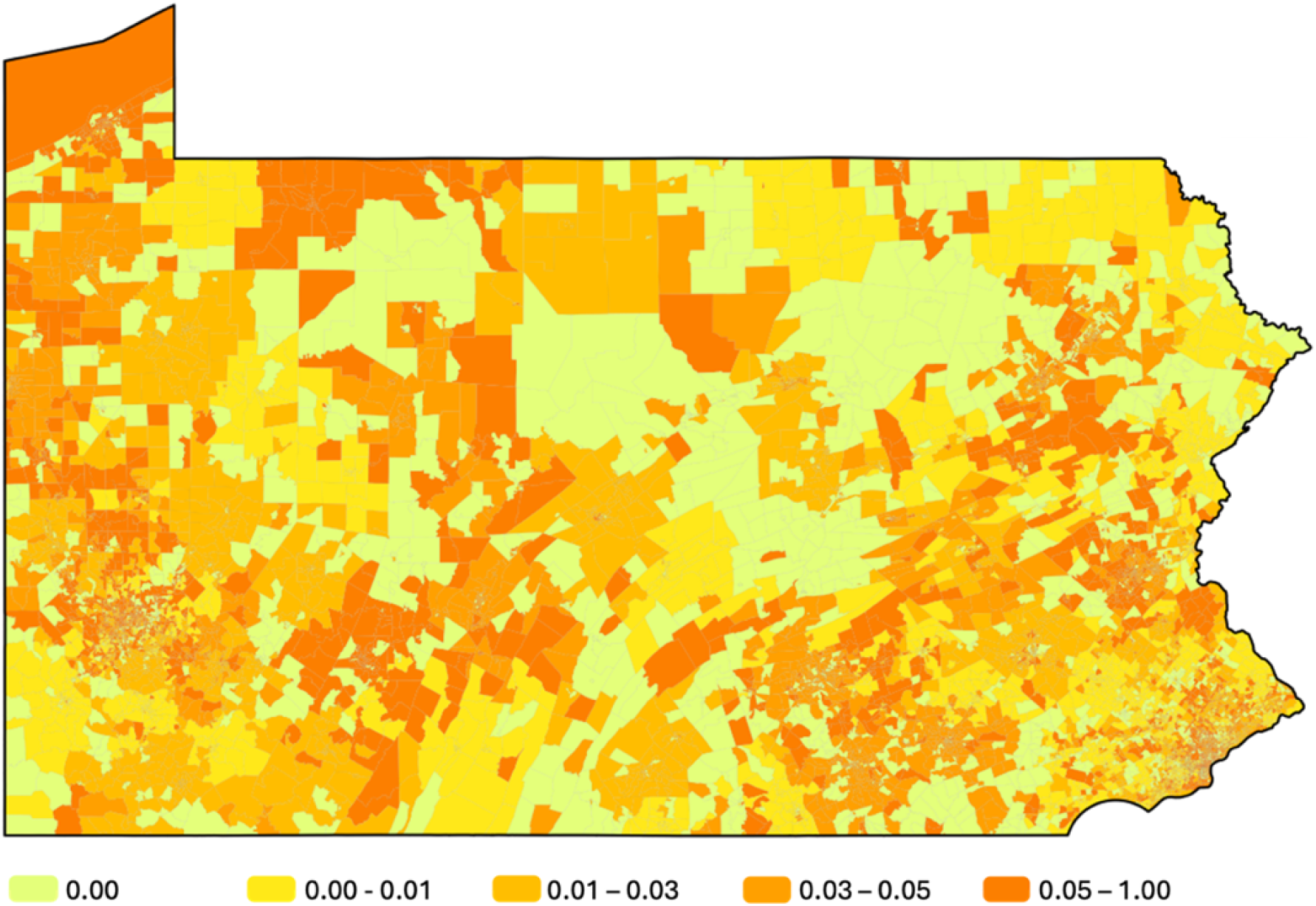
Accessibility map based on revised E2SFCA

## 5. Validation

Given the multifaceted nature of healthcare accessibility, validating spatial accessibility models is inherently challenging. Accessibility is shaped by a complex interplay of geographic, socioeconomic, behavioral, and institutional factors, many of which are not fully captured in spatial models. Considering this complexity, we validate our results through multiple complementary approaches, each offering insight into the realism and performance of the revised E2SFCA method.

### 5.1. Correlation with health outcomes

The first validation compares accessibility scores from both the traditional and revised E2SFCA models to public health outcomes, specifically the percentage of the population reporting poor or fair health across Pennsylvania counties. Because the health outcome data is available at the county level, we aggregated our block-group-level accessibility scores to match this spatial resolution. Figure 6 shows the association of accessibility models with county-level health status.

**Fig 6.**
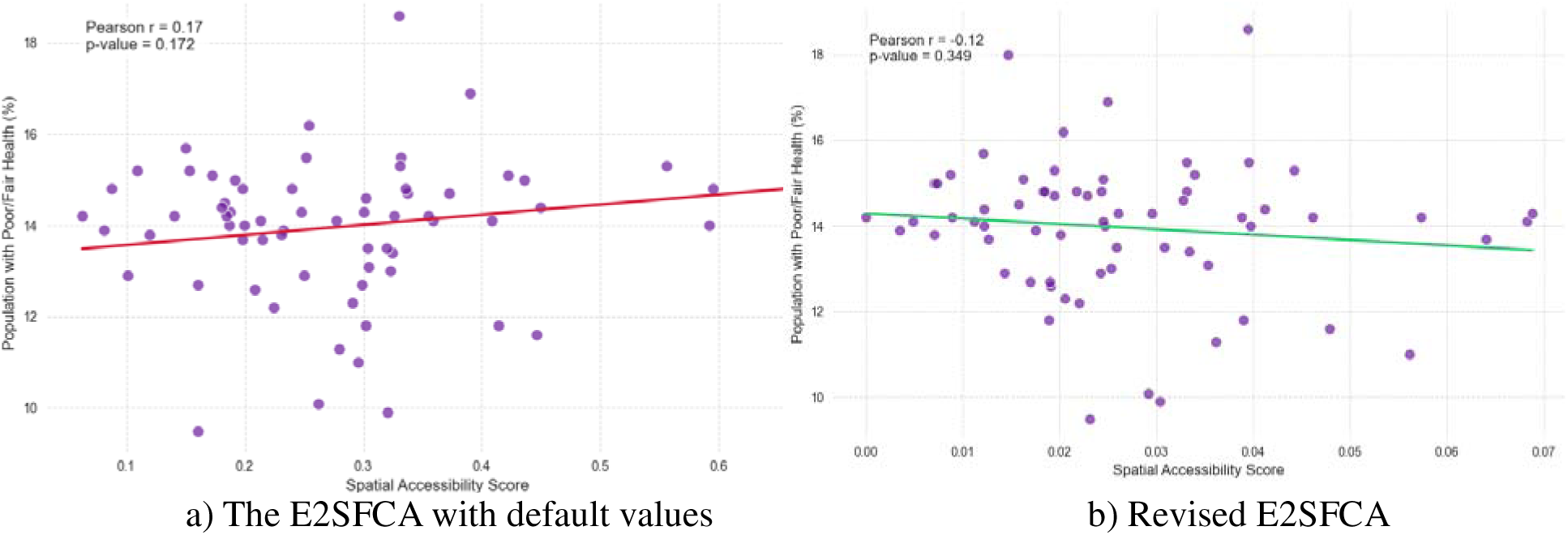
Comparative correlation of traditional and revised accessibility models with health outcomes

As shown in Figure 6a, the traditional E2SFCA method exhibits a weak positive correlation (r = 0.17, p = 0.172) with poor or fair health outcomes, suggesting higher accessibility is paradoxically associated with worse health, which contradicts real-world expectations. In contrast, the revised method (Figure 6b) shows a slightly negative correlation (r = –0.12, p = 0.35), which aligns more plausibly with the expectation that improved accessibility is associated with better health. While neither result is statistically significant, the revised model demonstrates a directionally more realistic relationship.

### 5.2. Correlation with household income

The second validation compares accessibility scores to median household income, under the premise that wealthier populations typically enjoy better spatial access to healthcare (Guo et al., 2022; Moscelli et al., 2018). Figure 7 represents comparative relationship between accessibility models and household income.

**Fig 7.**
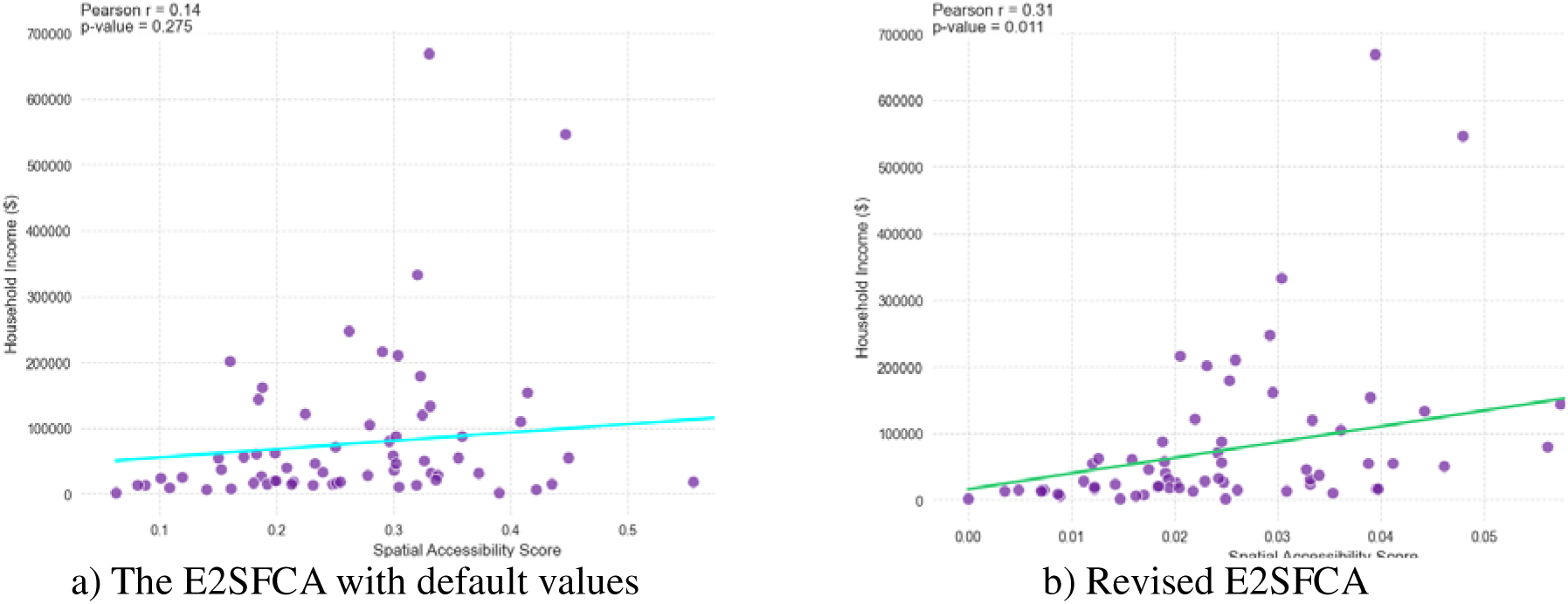
Accessibility and income: validating spatial models against socioeconomic indicators

As shown in Figure 7a, the traditional E2SFCA method yields a weak, non-significant correlation (r = 0.14, p = 0.27). However, the revised model (Figure 7b) shows a stronger and significant correlation (r = 0.31, p = 0.011), indicating that the revised scores better reflect socioeconomic patterns known to affect access.

### 5.3. Distributional equity: Gini Index

Finally, we assess equity in accessibility using the Gini index, which measures inequality in the distribution of healthcare access. The revised model reveals a more unequal landscape, with a Gini coefficient of 0.63, compared to 0.31 under the traditional E2SFCA approach. This substantial difference indicates that the revised method captures disparities that are obscured in conventional models (Chen et al., 2024).

To further illustrate this inequality, we calculated the cumulative distribution of accessibility and found that the top 19% of the population, ranked by access scores, accounts for 63% of total healthcare accessibility. This indicates a disproportionate concentration of access among a relatively small segment of the population. The revised model’s higher Gini index thus reflects entrenched inequities in spatial access, emphasizing the importance of moving beyond uniform assumptions to uncover the true extent of access inequality.

## 6. Limitations

While this study advances the E2SFCA framework by incorporating large-scale human mobility data, it is subject to several limitations related to both conceptual scope and data constraints.

### 6.1 Conceptual limitations

First, while the revised E2SFCA model improves upon traditional assumptions by leveraging empirical mobility patterns, it does not fully address several non-spatial factors that critically shape healthcare access. These include financial and insurance-related barriers, linguistic or cultural compatibility, healthcare needs varying by population subgroups, and system-level constraints such as provider capacity, appointment availability, or wait times. Moreover, like most spatial models, the analysis assumes that individuals have perfect knowledge of all providers within their catchment area and base decisions primarily on distance, which oversimplifies the complexity of healthcare-seeking behavior. Although observed mobility patterns implicitly capture some aspects of patient preference, such as the tendency to bypass nearby facilities, this approach does not fully account for the nuanced reasons behind such choices. Modeling these dimensions would require detailed behavioral, demographic, and institutional data beyond the scope of this study.

Additionally, the model is cross-sectional in nature and does not account for temporal dynamics in healthcare accessibility. While the mobility data offers some time granularity, the supply-side data such as hospital capacity is static. This limits the ability to assess how accessibility changes in response to factors like seasonal demand surges, temporary hospital closures, or staffing fluctuations.

### 6.2 Data limitations

The SafeGraph mobility data, while extensive, has inherent limitations. It represents an anonymized panel of mobile devices covering approximately 7.5% of the population (Li et al., 2024) and the sampling is uneven. Under-sampling is particularly notable among Hispanic, low-income, and low-education populations, which introduces potential biases in estimating true accessibility patterns. Furthermore, privacy protections and data aggregation result in the loss of granularity, as routes are not traceable and data are reported as flows or counts rather than individual-level trips. These limitations reduce the ability to infer exact travel paths or identify multimodal transport use, which may be especially relevant for underserved populations.

## 7. Conclusion

This study enhances how we measure healthcare accessibility by bridging spatial theory with the complexities of real-world human behavior. By integrating human mobility data into the E2SFCA framework, we uncover inequities that traditional models fail to detect. A key contribution of this work is its ability to enhance the empirical validity of the E2SFCA model by improving its alignment with real-world patterns without adding the complexity of new behavioral or demographic parameters. This balance of accuracy and simplicity makes the revised model particularly appealing for policymakers seeking actionable tools.

Compared to the default E2SFCA, the revised model demonstrates stronger correlations with public health outcomes and household income, also reveals deeper disparities through a substantially higher Gini index. These results clearly highlight how healthcare is uneven and disconnected across different areas. Although this study does not explicitly integrate variables such as hospital popularity or individual patient preferences, it shows that these factors are implicitly captured through mobility-informed decay functions and dynamic catchments. In doing so, the revised model elevates patterns of patient choice and institutional draw that are otherwise invisible to traditional frameworks.

Looking ahead, this work offers insights into more dynamic, responsive models of healthcare accessibility where accessibility scores adapt in real time to flu outbreaks, emergency events, or temporary hospital closures. While data limitations such as sampling bias in mobility datasets remain, the findings suggest that spatial accessibility models should evolve from static approximations into living systems that reflect the fluidity of human lives and the shifting realities of care infrastructure.

## Supporting information

Data and Results

## Data Availability

All data produced in the present study are available upon reasonable request to the authors.

https://doi.org/10.6084/m9.figshare.29856941

https://doi.org/10.6084/m9.figshare.29856926

## Funding

This work was supported by the National Institutes of Health [grant numbers R21MD018666 and R01AI174892].

## Notes

### Competing Interest Statement

The authors have declared no competing interest.

